# Fatigue Links Sociodemographic Risk to Pain Intensity and Spread in Two Surgical Cohorts

**DOI:** 10.64898/2026.02.02.26345387

**Authors:** Michael Sun, Briha Ansari, Daniel J. Clauw, Richard E. Harris, Kathleen A. Sluka, Chelsea M. Kaplan, Chad M. Brummet, Martin A. Lindquist, Tor D. Wager, The A2CPS Consortium

## Abstract

Why some surgical patients develop pain that extends beyond the original site of injury while others do not remains poorly understood. The recently proposed Risk of Pain Spread (ROPS) framework integrates multiple psychosocial risk factors associated with widespread pain, but its generalizability to surgical populations has not been established. Using two large pre-surgical cohorts of participants undergoing total knee arthroplasty or thoracic surgery, we evaluated the generalizability of the ROPS framework and examined relationships among psychosocial and sociodemographic factors, surgical-site pain, nonsurgical-site pain, and widespread pain using complementary latent-variable, regression, Bayesian network, and mediation analyses. Across cohorts and analytical approaches, fatigue emerged as the most consistent correlate of pain outcomes, independent of other measured risk factors. Fatigue also occupied a central position within the conditional dependency structure of psychosocial and pain variables and accounted for substantial shared variance linking sleep disturbance, depressive symptoms, neuroticism, adverse childhood experiences, and socioeconomic disadvantage with pain intensity. Nonsurgical-site pain consistently occupied a position linking pain intensity with widespread pain across complementary analyses. Together, these findings externally validate the ROPS framework in clinically relevant surgical populations and identify fatigue as a central correlate linking biopsychosocial vulnerability with pain burden.

**Author Note:** This work was funded by an administrative supplement from the National Institutes of Health as part of grant 5U54DA049110-04 (FOA RFA-RM-18-031). Data was provided by the A2CPS Consortium funded by the National Institutes of Health (NIH) Common Fund, which is managed by the Office of the Director (OD)/ Office of Strategic Coordination (OSC). Consortium components and their associated funding sources include Clinical Coordinating Center (U 24NS112873), Data Integration and Resource Center (U54DA049110), Omics Data Generation Centers (U54DA049116, U54DA049115, U54DA049113), Multi-site Clinical Center 1 (MCC1) (UM1NS112874), and Multi-site Clinical Center 2 (MCC2) (UM1NS118922).

**Conflicts of Interest:** Chad Brummett serves as a consultant for Vertex Pharmaceuticals and Merck Pharmaceuticals and provides expert medicolegal testimony. Tor Wager serves on the scientific advisory board of Curable Health. All other authors declare no competing interests.

## Introduction

Pain is a multidimensional experience with two core clinical features: its intensity and its tendency to spread beyond the initial site of injury. Pain intensity remains the primary focus of clinical care and research, yet widespread pain is often harder to predict, explain, and treat, and it carries greater disability, poorer quality of life, delayed recovery, and worse postoperative outcomes [16,19,33]. Understanding the mechanisms of pain spread is therefore an important challenge, particularly in surgical populations where tissue injury and recovery are relatively well characterized.

Numerous psychosocial and behavioral factors—depression, anxiety, fatigue, sleep disturbance, obesity, maladaptive coping, and life stress—are associated with greater pain severity and poorer postoperative recovery [5,11,13,15,16,23,24,29]. These characteristics rarely occur in isolation, however, and their overlap makes it difficult to separate independent contributors from downstream consequences of pain or shared expressions of broader biopsychosocial vulnerability. Most prior studies examine individual factors separately and focus on pain intensity, leaving open how multiple psychosocial factors jointly relate to widespread pain—which likely reflects interactions among neurobiological, psychological, and systemic processes rather than any single mechanism [2,8,18]. Pain catastrophizing, for example, is associated with greater acute postoperative pain [12,21], and genetic and epigenetic susceptibility may influence progression from localized to widespread pain [27].

Recent population-based research has begun to integrate these factors into unified risk frameworks. Using more than 500,000 UK Biobank participants, Tanguay-Sabourin and colleagues identified six modifiable factors—body mass index, fatigue, depression, neuroticism, sleep disturbance, and life stress—that together comprise the Risk of Pain Spread (ROPS) framework and predict the presence and extent of widespread pain [30]. Rather than treating these factors independently, ROPS conceptualizes them as an interconnected profile of modifiable risk, within which fatigue is among the strongest predictors and may represent a proximal manifestation through which cumulative vulnerability is expressed [5,30].

Whether this community-derived framework generalizes to clinical surgical populations remains unknown. Compared with community volunteers, pre-surgical patients present with established, clinically characterized pain—such as an osteoarthritic knee scheduled for arthroplasty—greater distress, heterogeneous pain mechanisms, and imminent tissue injury, any of which could alter these relationships [16,26]. The UK Biobank data used to derive ROPS also lacked validated, localized pain sites and did not model how the risk factors are organized across multiple pain outcomes.

We therefore examined two independent pre-surgical cohorts from the Acute to Chronic Pain Signatures (A2CPS) Consortium [3,26] with three aims: to test whether the ROPS framework generalizes across surgical cohorts; to determine whether its factors relate not only to widespread pain but also to surgical-site and non-surgical-site pain intensity; and to characterize how the factors are organized in relation to one another, using complementary latent-variable, regression, Bayesian network, and mediation analyses. Characterizing this organization may reveal central, potentially modifiable targets that are not apparent when risk factors are studied in isolation.

## Methods

### Experimental Design

We analyzed baseline data from two independent pre-surgical cohorts drawn from the A2CPS Consortium – Data Release 2.0.0, which included participants preparing for total knee arthroplasty (TKA) and participants preparing for thoracic surgery. These cohorts differ in surgical indication, pain burden, and demographic composition, allowing us to examine whether associations between psychosocial risk factors and pain outcomes generalize across distinct pre-surgical contexts. We focused on three pain outcomes assessed prior to surgery: worst intensity of surgical-site pain (SSP), worst intensity of non-surgical-site pain (NSSP), and chronic widespread pain, defined as the number of body regions with moderate or greater pain persisting for at least six months. Predictors included sociodemographic variables (age, sex, marital status, income, and education) and psychosocial and behavioral factors corresponding to the Risk of Pain Spread (ROPS) framework, including body mass index, fatigue, depression, neuroticism, sleep disturbance, and adverse childhood experiences. Because no single statistical framework can fully characterize complex multivariable relationships, we employed complementary latent-variable, regression, mediation, and network analyses that address distinct but related questions regarding association structure, conditional dependence, and statistical directionality. We first estimated latent-variable and regression-based models to characterize univariate and multivariable associations. We then used network and mediation analyses to assess patterns of statistical dependence among variables and to identify factors occupying central positions within association networks. We conducted all analyses separately within each cohort and compared results across cohorts to assess robustness and cohort sensitivity. We integrated these complementary analyses to identify convergent patterns that generalized across cohorts.

### Baseline A2CPS Assessment and Sample Inclusion Criteria

Data collection involved harmonized, standardized protocols across study sites, including self-report questionnaires, clinical evaluations, and behavioral measures. The shared and cohort-specific eligibility criteria can be found in Table 1 of Berardi, et al., 2022 [3]. Participants were not excluded for prior chronic pain conditions, including fibromyalgia, neuropathic pain, peripheral neuropathy, low back pain, mental health conditions, or prior surgery > 3 months previously; or based on prior or baseline pain medication use, including opioids, to maximize generalizability. A full Schedule of Activities can be found in Supplementary Table S3 of Berardi et al., 2022 [3]. Inclusion criteria for the present analysis were complete baseline phenotyping data and availability of key sociodemographic and ROPS variables. Participants were excluded if they withdrew consent, or were deemed ineligible by site investigators.

**Table 1.** Participant characteristics and baseline pain profiles by surgical cohort. Baseline sociodemographic characteristics, Risk of Pain Spread (ROPS) factors, and pain measures for total knee arthroplasty (TKA) and thoracic surgery cohorts. Continuous variables are reported as mean (SD) unless otherwise noted; categorical variables are reported as percentages or modal category. Group differences were assessed using Welch’s t tests for continuous variables and χ² tests for categorical variables. Pain duration is reported as median and mean (SD). widespread pain reflects the number of body sites with pain intensity ≥3 lasting ≥3 months.

|  | TKA | Thoracic | Test Statistic | p-value |
| --- | --- | --- | --- | --- |
| N | 1043 | 358 |  |  |
| **Demographics** |  |  |  |  |
| Age Mean (SD) | **65.2 (8.5)** | 59.8 (13) | $t(449.45) = 7.22$ | $2.27e-12^{***}$ |
| Sex (Mode) | Female | Female | $\chi^2(1) = 5.78$ | $0.02^*$ |
| % Male | 36.5 | **44.1** |  |  |
| % Female | **62.2** | 55.3 |  |  |
| Marital Status (Mode) | Married | Married | $\chi^2(6) = 1.79$ | 0.94 |
| Income Level (Mode) | **100k-149.9k** | 50k-74.9k | $\chi^2(10) = 50.69$ | $2e-7^{***}$ |
| Education Level (Mode) | **Postgrad** | High School | $\chi^2(6) = 53.19$ | $1.07e-9^{***}$ |
| Employment Status (Mode) | Not Employed | Not Employed | $\chi^2(3) = 8.49$ | $0.04^*$ |
| Applied for Disability (Mode) | No | No |  |  |
| % Applied for Disability | 11.1 | **15.9** | $\chi^2(2) = 8.42$ | $0.02^*$ |
| **Risk of Pain Spread (ROPS)** |  |  |  |  |
| Adverse Childhood Events (ACEs) | 1.51 | 1.67 | $t(537.65) = 1.15$ | 0.25 |
| Body Mass Index (BMI) | **31.25** | 29.33 | $t(470.58) = 4.83$ | $1.88e-6^{***}$ |
| Depression (PHQ) | 4.34 | **5.01** | $t(523.93) = 2.16$ | $.03^*$ |
| Fatigue | **17.53** | 16.68 | $t(521.88) = -2.04$ | $.04^*$ |
| Neuroticism (BIG5) | 8.80 | **9.73** | $t(510.87) = 3.80$ | $1.62e-4^{***}$ |
| Sleep Problems (PROMIS) | 2.87 | 2.76 | $t(539.4) = -1.73$ | $.08^\dagger$ |
| **Pain** |  |  |  |  |
| Pain Duration Median | **36 months** | 1 month | $t(654.90) = 6.02$ | $2.93e-9^{***}$ |
| Mean (SD) | **71.4 (173.8)** | 17.1 (120.4) |  |  |
| Surgical Site Pain Intensity (SSP) | **6.16** | 0.99 | $t(679.12) = -35.67$ | $8.39e-158^{***}$ |
| Nonsurgical Site Pain Intensity (NSSP) | **4.10** | 2.18 | $t(679.12) = -35.67$ | $1.82e-23^{***}$ |
| Widespread Pain | **1.51** | 1.11 | $t(679.12) = -3.65$ | $2.85e-4^{***}$ |
| Widespread Pain (Modal Body Sites) | 0 | 0 | $\chi^2(6) = 7.29$ | $0.03^*$ |
| Widespread Pain (%0 Body Sites) | 41.6 | **69.3** |  |  |
| Widespread Pain (%1 Body Sites) | **24.9** | 8.7 |  |  |
| Widespread Pain (%2 Body Sites) | **17.5** | 8.1 |  |  |
| Widespread Pain (%3 Body Sites) | **8.5** | 5 |  |  |
| Widespread Pain (%4+ Body Sites) | 7.4 | **8.9** |  |  |

$^\dagger p < .10$ , $^* p < 0.05$ , $^{**} p < 0.01$ and $^{***} p < .001$ .

With the sparse sample size across multiple levels of each demographic variable, we dropped the Intersex/Unknown (n = 15) levels of the sex variable, dropped the Unknown education level (n = 32), aggregated the marital status variable to Non-Married, Married/Domestic Partner, and dropped the Unknown marital status (n = 31) level, and converted the income status variable into a continuous one by setting each participant’s income status to the median of their reported income range.

### Pain Intensity and Spread – Brief Pain Inventory and Michigan Body Map

Pain was assessed using the Brief Pain Inventory (BPI; [9]) and the Michigan Body Map (MBM; [4]. The BPI was modified to assess the worst pain intensity at the pre-surgical site over the past 24 hours with anchors of 0 (no pain) to 10 (worst pain imaginable). The Michigan Body Map was used to record the number of painful body regions with persistent pain over the past 3 months (at baseline).

Each participant in the Thoracic surgery and TKA cohorts was categorized into pain subtypes using the BPI and MBM. Individuals were classified as either having No Chronic Pain or Chronic Pain (any pain duration ≥ 3 months as measured by the MBM). Within the No Chronic Pain category, subtypes included: no pain reported at all (No Pain), no pain in the last 24 hours (as measured by the BPI; No Pain < 24hrs), minimal pain in the last 24 hours (pain intensity ≤ 3; Minimal Pain < 24hrs) or some acute pain in the last 24 hours (Acute Pain < 24hrs). Those in the Chronic Pain category were further divided into two subtypes: Low Chronic Pain (pain intensity ≤ 3 and any pain duration ≥ 3 months) and Moderate/High Intensity Chronic Pain (pain intensity > 3 and pain duration ≥ 3 months in at least one body site).

### ACEs

The Adverse Childhood Experience questionnaire (ACE [10]) is a 10-item retrospective assessment of childhood (age 0– 18 years) abuse, neglect, and household dysfunction. Each item is scored dichotomously (0 = absent, 1 = present), and summed to an ACEs total score ranging from 0 to 10. Higher scores reflect greater cumulative exposure to early adversity.

### Neuroticism

The Neuroticism (Negative Emotionality) subscale was derived from the Big Five Inventory−2 short form (BFI-2-S [28], comprising of four items: “Is temperamental, gets emotional easily,” and reverse codings of “Is emotionally stable, not easily upset,” “Is relaxed, handles stress well”, and “Is persistent, works until the task is finished.” Participants rated each item according to their level of agreement from 1 (disagree strongly) to 5 (agree strongly). Responses to these items were averaged to yield a mean Neuroticism score, scored out of 20, with higher values indicating greater emotional instability and sensitivity to stress.

### Depression

Depression was assessed using the Patient Health Questionnaire-8 (PHQ-8 [17], which included questions about participant experiences during the past 2 weeks, scored from 0 (“not at all”) to 3 (“nearly every day”). Higher total scores represented elevated levels of depression.

### Sleep Disturbance

Sleep disturbance is assessed using the Patient Reported Outcomes Measurement Information System (PROMIS) Short Form v1.0 – Sleep Disturbance 6a measure [34]. Subjects rate their sleep quality and difficulty falling asleep over the past 14 days on a scale of 1 (not at all) to 5 (very much) with higher scores indicating greater sleep disturbance.

### Fatigue

Fatigue was assessed using the PROMIS Short Form v1.0 – Fatigue 7a [1]. This seven-item measure has Likert-scale responses ranging from 1 (never) to 5 (always) with higher scores indicating greater fatigue.

### Latent variable analyses

We used structural equation modeling (SEM) with lavaan (*sem()*) to define a latent Risk of Pain Spread (ROPS) construct and examine its association with SSP, NSSP, and widespread pain. The latent ROPS factor was specified using the six observed ROPS predictors. All continuous variables were standardized prior to modeling. Models adjusted for demographic covariates, accounted for correlated pain outcomes, and handled missing data using full information maximum likelihood with robust estimation. Each outcome was modeled simultaneously, with residual correlations specified between pain outcomes to account for shared variance not explained by the latent factor. Cohort differences were evaluated using multi-group SEM and nested model comparisons.

### Correlogram, dendrogram, and tanglegram analyses

Pairwise correlations among predictors and outcomes were computed using Spearman’s coefficients and visualized with correlograms. Hierarchical clustering was applied to correlation matrices, and dendrograms were compared across methods using tanglegrams to evaluate cluster stability. Correlograms were generated separately for each cohort and variables were clustered using Ward’s minimum variance method (Ward.D2). To compare the resulting dendrograms across cohorts, we used tanglegrams, where colors indicated cluster membership. We computed two complementary metrics: entanglement, which quantifies how well two trees can be aligned without crossing edges (0 = identical, 1 = maximally tangled), and Baker’s γ correlation, which assesses concordance of ultrametric distances between the trees (−1 to 1). Significance of Baker’s γ was evaluated using 5,000 permutations of the dendrogram labels to generate a null distribution.

### Single-variable and multivariable regression analyses

Associations between predictors and pain phenotypes were examined using generalized linear models. Ordinary Least Squares (OLS) multiple regression models were first fit within the Thoracic surgery and TKA participant cohorts to assess model assumptions. Residuals and leverage values were extracted from each model to evaluate multicollinearity (Variance Inflation Factor – VIF), heteroskedasticity (Breusch–Pagan χ² – BP χ²), non-normality (Shapiro–Wilk W – SW-W), influence (Cook’s D), and a global omnibus test of skewness, kurtosis, and functional form (Global Validation of Linear Model Assumptions; using the *gvlma* package in R). Regression diagnostics indicated distinct patterns of model assumption violations across cohorts and outcomes. In both cohorts, VIFs showed no multicollinearity issues. The SSP model showed the most severe assumption breaches, with heteroscedasticity (BP χ² = 43.66, p < .001), non-normality (SW-W p < .001), and violations of skewness and kurtosis (ps < .05), alongside several influential observations. For NSSP, both heteroscedasticity (BP χ² = 31.58, p = .005) and non-normal residuals (SW-W p < .001) were present, with multiple influential cases; global tests again suggested skewness departures. For spread, residual variance was stable (BP χ² = 17.41, p = .18), though residuals deviated from normality (SW-W p = .003), and several observations were highly influential. Global validation tests further flagged skewness violations. In the TKA cohort, the SSP model showed stable variance (BP χ² = 12.66, p = .70), but residuals were non-normal (SW-W p < .001) and influenced by a small number of high-leverage points. By contrast, for NSSP, residuals were heteroscedastic (BP χ² = 30.84, p = .014) as well as heavy-tailed (SW-W p = .004), with kurtosis flagged in global tests. For widespread pain, residuals were also heteroscedastic (BP χ² = 36.20, p = .003), non-normal (SW-W p < .001), and flagged for skewness, kurtosis, and link function misspecification (ps < .05).

Given consistent OLS violations across cohorts involving non-normal residuals, influential cases, and global test failures related to skewness and heavy tails, OLS regression models were performed on log-transformed (log(1+y)) outcomes on the worst pain ratings for both SSP and NSSP. Coefficients were back-transformed so that for each 1-unit increase in a predictor we report the percent change in worst pain: Δ%=(−1)×100%. Quasipoisson regression (log link) was used for the count of spread given its negative skew. We likewise report percent change using (−1)×100%, which is equivalent to (Incidence Rate Ratio IRR−1)×100%. McFadden R^2^ was computed for quasipoisson models as a proxy for variance explaining spread. First, “single-predictor” regression models were fit where each outcome was regressed upon by a single predictor, cohort (effects coded centered at 0 with Thoracic set as –1, and TKA as 1), and the predictor x cohort interaction. This was followed by multiple regression models where all predictors were fit simultaneously in an independent model for each cohort. Multiple regression models were ten-fold cross-validated to estimate the Variable Importance (VI) out of 100 of each predictor’s inclusion in each model. Employment status and application for disability were not tested as predictors because of their likelihood of codetermination with pain symptom outcomes. Predictor variables were explicitly coded as either discrete (categorical) or continuous (numeric) to permit estimation under a conditional Gaussian framework. Categorical demographic variables (e.g., sex, marital status, education) were treated as factors, while continuous measures (e.g., age, income, ACEs, BMI, neuroticism, sleep quality, fatigue, depression, spread and log-transformed SSP and NSSP) were modeled as numeric.

### Network analyses

Bayesian network structure learning was selected because it estimates conditional dependencies among variables while allowing comparison of competing directional hypotheses under explicit structural constraints, thereby generating probabilistic hypotheses regarding statistical directionality without assuming a predetermined recursive regression ordering. To assess conditional dependencies between predictors and outcomes, we estimated partial correlation networks using the graphical least absolute shrinkage and selection operator (GLASSO) with extended Bayesian information criterion (EBIC) model selection to improve model sparsity. Network centrality indices (strength, closeness, betweenness) were computed to identify influential variables. Cases with missing data were excluded listwise. We applied a score-based *tabu* search algorithm (*bnlearn* v.4.9 [25] using the Bayesian Information Criterion for conditional Gaussian networks (BIC-cg) as the model selection score. To enforce valid conditional Gaussian models, edges from continuous to discrete variables were blacklisted, preventing Gaussian parents of categorical children. Additional blacklisted constraints were incorporated where theoretically justified e.g., demographic variables set as exogenous, pain outcome variables spread, SSP, and NSSP were set as endogenous, and endogenous edges to household income, age, BMI, ACEs, and neuroticism from sleep quality, fatigue, and depression were blacklisted. To interpret the networks, computed centrality metrics included indegree, outdegree, total degree, betweenness centrality, closeness, authority, pagerank centrality, eigenvector centrality, hubscore, and distance to outcome.

### Counterfactual network analyses

To evaluate competing hypotheses regarding statistical directionality directional hypotheses between fatigue and pain, we constructed four model classes: Fatigue→Pain (F2P), Pain→Fatigue (P2F), Unconstrained (UNC), and Fatigue-as-Outcome (FOUT) by restricting or permitted specific edges using whitelists (required directions) and blacklists (forbidden directions). All models enforced plausible constraints preventing reverse causation into trait-like variables (e.g., age, sex, income) and disallowing continuous→categorical edges to satisfy the conditional Gaussian (CG) model assumptions. Each model class was fit separately within the Thoracic surgery and TKA cohorts. To assess relative evidence for each directional hypothesis, BIC-cg scores were compared between models, where higher values indicate better model fit after penalizing complexity. We summarized model fit across classes by computing ΔBIC relative to the best model for each cohort and ranked models accordingly. To evaluate the robustness of key directional edges, we repeated structure learning using nonparametric bootstrapping (R = 5,000) with the same whitelist/blacklist constraints and computed arc strength and direction probabilities using *boot.strength*.

### Mediation analyses

Mediation models were estimated using the mediation and lavaan R packages [14]. These analyses were performed to complement the Bayesian network analyses by quantifying indirect statistical associations and should not be interpreted as evidence of causality. Average Indirect Effects (AIE), Average Direct Effects (ADE), and Proportion Mediated (PM) were quantified, with 5,000 bootstrap samples used to generate bias-corrected 95% confidence intervals. Mediation pathways tested whether ROPS predictors influenced spread indirectly via fatigue or NSSP.

For continuous outcomes (SSP and NSSP), we fit observed-variable path models using SEM with lavaan (*sem()*), specifying two regression equations per mediation:

1. M ∼ X + covariates (paths “a”)
2. Y ∼ M (path “b”) + X (paths “c′”) + covariates.

We used quasipoisson models for spread and a linear model for the mediator and effects are reported on the expected count scale. Following the counterfactual mediation framework [22], for DAG-indicated pathways within each cohort we estimated:

1. natural indirect effects (AIE = E[Y(x, M(x)) − Y(x, M(x′))]
2. direct effects (ADE = E[Y(x, M(x′)) − Y(x′, M(x′)]
3. total effects (Total Effect: TE = AIE + ADE)
4. Proportion mediated (at control n_0_): PM = AIE / TE

95% confidence intervals (CIs) were obtained via nonparametric bootstrap with bias-corrected and accelerated (BCa) intervals. We targeted 5,000 resamples. Mediation effects (AIE, ADE, TE) were estimated with the mediation package (*mediate()*).

### Statistical Analysis

All analyses were conducted in R (version 4.5.1) using two-sided tests with α = 0.05. Continuous predictors were standardized prior to analysis. Effect sizes and 95% confidence intervals are reported alongside p values where appropriate. Additional details regarding each analytical approach are provided in the preceding sections.

### Ethical approval

All procedures were approved by a single Institutional Review Board (IRB) at the University of Iowa. All participants provided written informed consent in accordance with the Declaration of Helsinki.

## Data and Materials Availability

All data provided from the A2CPS consortium are available to other investigators online upon permission granted by https://a2cps.org/researchers/accessing-our-data/. Detailed analysis code and annotation will be made publicly available at GitHub (https://github.com/canlab and https://github.com/a2cps).

## Results

### Participant Characteristics

Participant characteristics (Fig. 1, 2) are summarized in Table 1. Females comprised 55% of the Thoracic surgery cohort and 62% of the TKA cohort, reflecting a significantly different female-to-male ratio between cohorts (χ²(1) = 5.78, p = 0.02). TKA participants were older (mean ± SD: 65.20 ± 8.50 years vs. 59.80 ± 13 years; t(449.45) = 7.22, p = 2.27e-12), more educated (modal educational attainment: Postgrad vs. High School; χ²(6) = 53.19, p = 1.07e-09), more likely to be employed (χ²(3) = 8.49, p = 0.04), more likely to be in a higher income bracket (modal income bracket: $100k-$149.9k vs. $50k-$74.9k; χ²(10) = 50.69, p = 2.00e-07), and less likely to have applied for disability (χ²(2) = 8.42, p = 0.02). No significant differences were observed in marital status between cohorts (χ²(6) = 1.79, p = 0.94).

**Figure 1.**
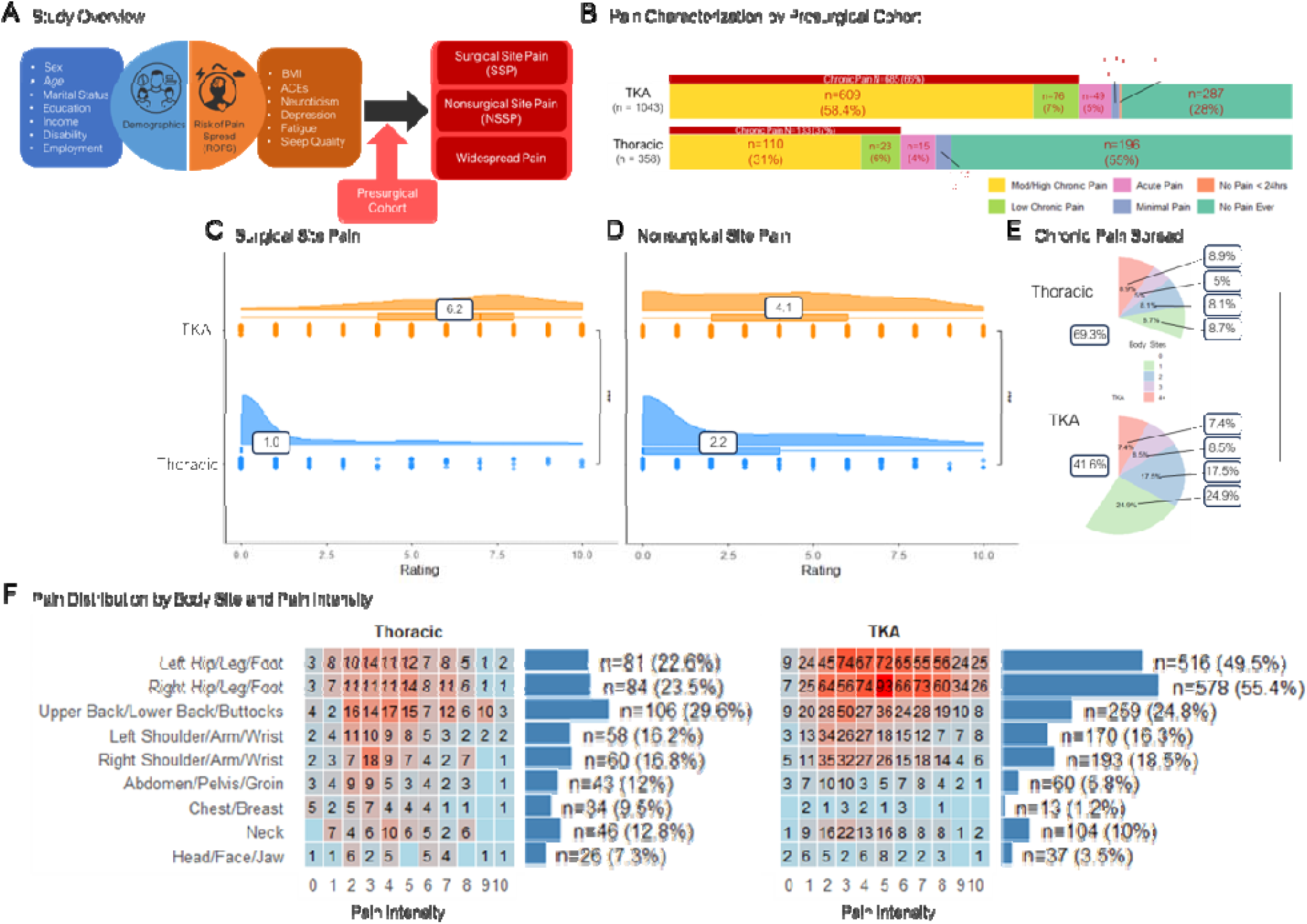
Study design and baseline pain characteristics across surgical cohorts. (A) Study overview and analytical framework. Presurgical pain outcomes—surgical-site pain intensity (SSP), non-surgical-site pain intensity (NSSP), and widespread pain—were modeled as a function of sociodemographic variables and analogues of Risk of Pain Spread (ROPS) predictors, with cohort (thoracic surgery vs. total knee arthroplasty [TKA]) examined as a potential moderator. (B) Distribution of pain subtypes across cohorts using the Michigan Body Map. Pain ratings ≥3 defined moderate-to-high pain. Pain was classified as chronic if reported to last ≥3 months. (C–E) Cohort distributions of presurgical pain outcomes. Shown are distributions of (C) SSP, (D) NSSP, and (E) widespread pain. TKA participants reported higher SSP and NSSP and a greater number of chronic pain sites among individuals with chronic pain. TKA participants were more likely than thoracic surgery participants to report one to three painful sites (χ²(4) = 96.69, p = 2.22 × 10 ¹), whereas rates of four or more painful sites (widespread pain) did not differ significantly between cohorts. (F) widespread pain by body site and intensity. Heatmap shows the number of participants reporting pain at each body site and intensity level (any duration), stratified by cohort.

Among ROPS factors (Fig. 2F-K), thoracic surgery participants scored higher on neuroticism (t(510.87) = 3.80, p < .001, *g*=0.25) and depression (t(507.4) = 3.27, p < .001, *g* = 0.21) than TKA participants. Conversely, TKA participants had higher BMI (31.25 vs. 29.33; t(470.58) = 4.83, p < .001, *g*=0.32), greater fatigue (t(521.88) = 2.04, p = .04, *g*=.13), and marginally more sleep problems (t(539.4) = –1.73, p = .08) than Thoracic participants.

**Figure 2.**
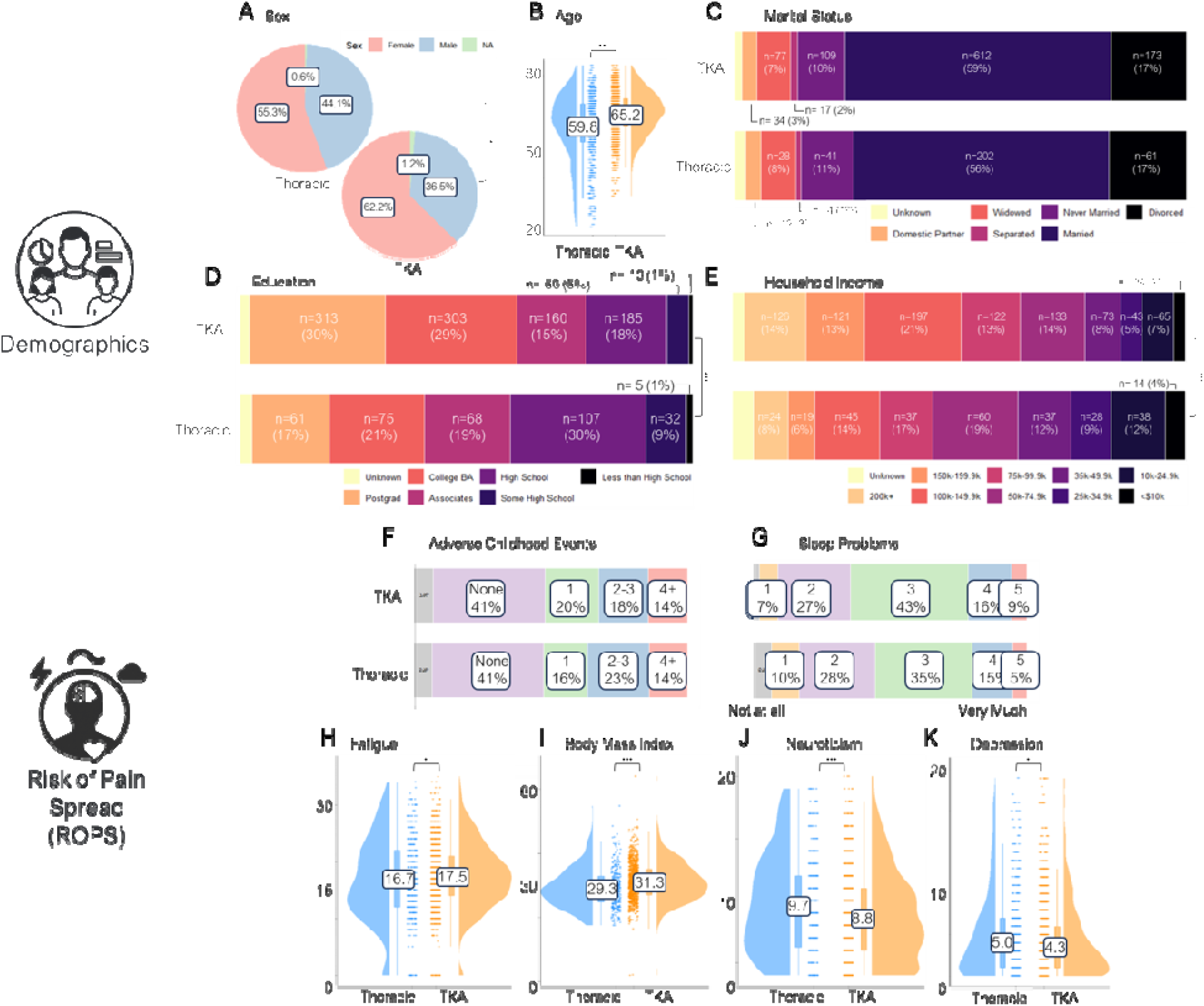
Sociodemographic and psychosocial differences across surgical cohorts. (A–E) Sociodemographic characteristics by cohort. The thoracic surgery and total knee arthroplasty (TKA) cohorts differed significantly in sex composition, age, education, and household income. Relative to the thoracic cohort, the TKA cohort was older, more highly educated, and reported higher income. (F–K) Risk of Pain Spread (ROPS)–related factors by cohort. The TKA cohort showed higher body mass index and greater fatigue, whereas the thoracic surgery cohort reported higher levels of neuroticism and depressive symptoms.

**Figure 3.**
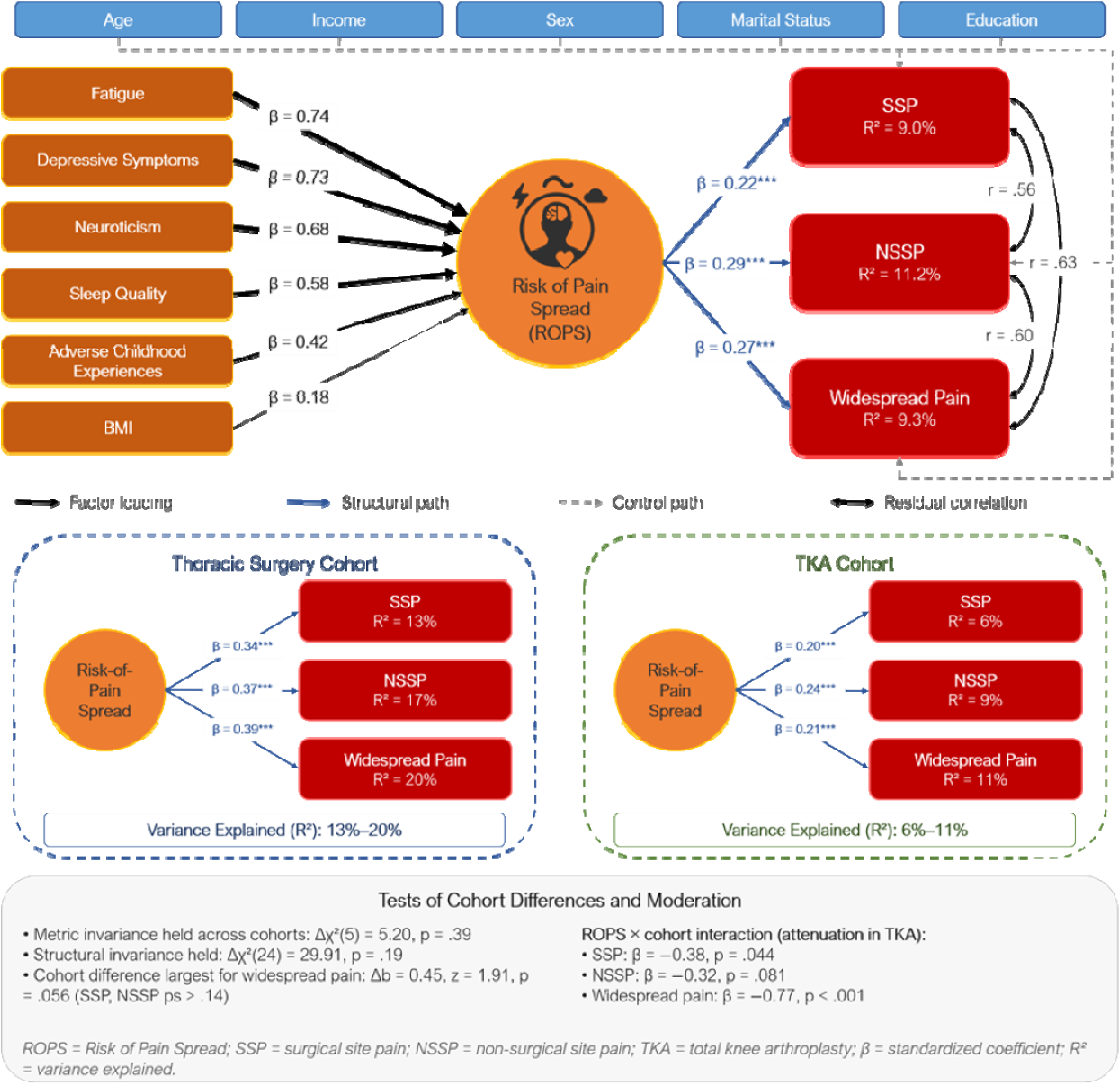
Structural equation model of the Risk of Pain Spread (ROPS) framework across surgical cohorts. Top: Measurement and structural model of the latent ROPS construct predicting surgical-site pain (SSP), non-surgical-site pain (NSSP), and widespread pain while adjusting for sociodemographic covariates. Bottom: Cohort-specific standardized structural path coefficients and explained variance (R²) for the thoracic surgery and total knee arthroplasty (TKA) cohorts. Measurement invariance and tests of cohort moderation are summarized below the models.

Using the Brief Pain Inventory-Short Form (BPI-SF) to assess worst pain intensity over the past 24 hours, TKA participants reported higher [pre-]surgical site pain (hereinafter SSP – mean ± SD: 6.16 ± 2.59 vs. 0.99 ± 2.18; t(679.12) = 35.67, p < 8.39e-16) and greater nonsurgical site pain (NSSP – 4.10 ± 2.93 vs. 2.18 ± 2.82; t(562.71) = 10.45, p < 1.82e-23) compared with Thoracic surgery participants. Widespread chronic pain (Fig. 1B, E) was defined as the number of body regions (out of nine) with pain intensity ≥ 3 sustained for ≥ 6 months using the Michigan Body Map. Individuals were categorized into one of five levels (0, 1, 2, 3, or ≥ 4 body sites in chronic pain). Individuals with TKA were more likely to report pain in one and two-to-three body sites (χ²(4) = 96.69, p = 2.22e-16), whereas the prevalence of four or more sites (i.e., widespread pain) did not differ significantly.

### Latent variable analysis associating ROPS with pain outcomes

We first asked whether the latent ROPS architecture identified in UK Biobank replicated in two independent presurgical cohorts. To test whether a latent Risk of Pain Spread (ROPS) construct predicts pain burden, we fit a multivariate structural equation model with a single latent ROPS factor predicting log-SSP, log-NSSP, and widespread pain, while adjusting for age, income, sex, marital status, and education. The ROPS latent factor was specified by the six ROPS indicators: adverse childhood experiences, body mass index, neuroticism, sleep quality, fatigue, and depressive symptoms.

### Measurement Model

All ROPS indicators loaded significantly on the ROPS latent construct (β: 0.18–0.78; *ps* < .001). Loadings were strongest for fatigue (β = 0.74), depressive symptoms (β = 0.73), and neuroticism (β = 0.68), with more modest contributions from sleep quality (β = 0.58), adverse childhood experiences (β = 0.42), and BMI (β = 0.18). Model-based reliability of the latent factor was acceptable (ω ≈ 0.61), indicating that the ROP construct captured shared variance across heterogeneous risk indicators.

### Structural associations with pain outcomes

In the full multivariate model, the latent ROPS factor significantly predicted all three pain outcomes after adjusting for demographic covariates. Higher ROPs scores were associated with greater SSP (β = 0.22, *p* < .001), NSSP (β = 0.29, *p* < .001), and widespread pain (β = 0.27, *p* < .001). The model explained 9.0% of the variance in SSP, 11.2% in NSSP, and 9.3% in widespread pain. Residual correlations among pain outcomes were moderate to large (all *ps* < .001), supporting the use of a multivariate outcome model.

### Cohort-specific effects

To evaluate whether the ROPS–pain associations differed by clinical cohort, we conducted a multi-group SEM comparing Thoracic surgery and TKA cohorts. Measurement invariance testing supported equivalence of factor loadings across cohorts (metric invariance: Δχ²(5) = 5.20, *p* = .39), indicating that the ROPs construct was psychometrically comparable. Constraining all structural paths to equality across cohorts did not significantly worsen model fit (Δχ²(24) = 29.91, *p* = .19), suggesting no global violation of structural invariance. Despite this, cohort-specific estimates revealed systematically stronger ROPS–pain associations in the Thoracic surgery cohort. In Thoracic participants, the latent ROPS factor showed large, standardized associations with SSP (β = 0.34), NSSP (β = 0.37), and widespread pain (β = 0.39), whereas corresponding effects in the TKA cohort were more modest (βs = 0.20–0.24 for severity and interference; β = 0.21 for widespread pain). The latent ROPS factor explained a greater proportion of variance in all outcomes in the Thoracic surgery cohort (R² range: 13–20%) than in the TKA cohort (R² range: 6–11%).

### Tests of cohort differences and moderation

Pairwise comparisons of cohort-specific path coefficients indicated that differences between cohorts were marginally different for widespread pain (Δb = 0.45, *z* = 1.91, *p* = .056), with smaller and non-significant differences for SSP and NSSP (*ps* > .14). To formally test moderation, regression models using latent ROPS factor scores revealed significant ROPS × cohort interactions, demonstrating attenuation of ROPS effects in the TKA cohort. Interaction effects were observed for SSP (β = −0.38, *p* = .044) and were strongest for widespread pain (β = −0.77, *p* < .001), with a marginally significant effect for NSSP (β = −0.32, *p* = .081).

### Tanglegram analysis

Hierarchical clustering of cohort-specific Spearman correlation matrices revealed highly similar higher-order organization of psychosocial and pain variables across the Thoracic surgery and TKA cohorts, despite substantial demographic and clinical differences (Supplementary Fig. S1). Fatigue was the most consistently positively correlated variable with SSP, NSSP, and widespread pain in both cohorts. SSP and NSSP, and NSSP and widespread pain, were strongly positively correlated in both cohorts, whereas SSP correlated with widespread pain only in the Thoracic surgery cohort. Bivariate correlations further showed that younger age was associated with higher SSP in both cohorts and higher NSSP in the TKA cohort, and that in the TKA cohort female sex was associated with higher NSSP, unmarried or unpartnered status with higher NSSP and widespread pain, and lower income with higher pain outcomes. Depression, neuroticism, sleep problems, and fatigue formed a robust ROPS cluster correlating with all pain outcomes in the TKA cohort; in the Thoracic cohort, neuroticism did not correlate with SSP, and depression, neuroticism, and sleep did not correlate with NSSP, although all correlated with widespread pain. ACEs and BMI correlated with NSSP and widespread pain in the TKA cohort but not the Thoracic cohort. Tanglegram analysis confirmed the similarity of the clustering structures across cohorts (Fig. 4; ε = 0.11; Baker’s γ = 0.49, p = .003): ROPS variables other than BMI clustered with the pain outcomes— forming a single cluster in the Thoracic cohort and bifurcating in the TKA cohort—while sociodemographic variables and BMI grouped separately in a residual space of low correlation with the ROPS and pain variables. Together, these descriptive clustering analyses indicate a largely conserved multivariable architecture across cohorts and motivated the subsequent Bayesian network analyses, which formally characterized the conditional dependency structure among these variables.

**Figure 4.**
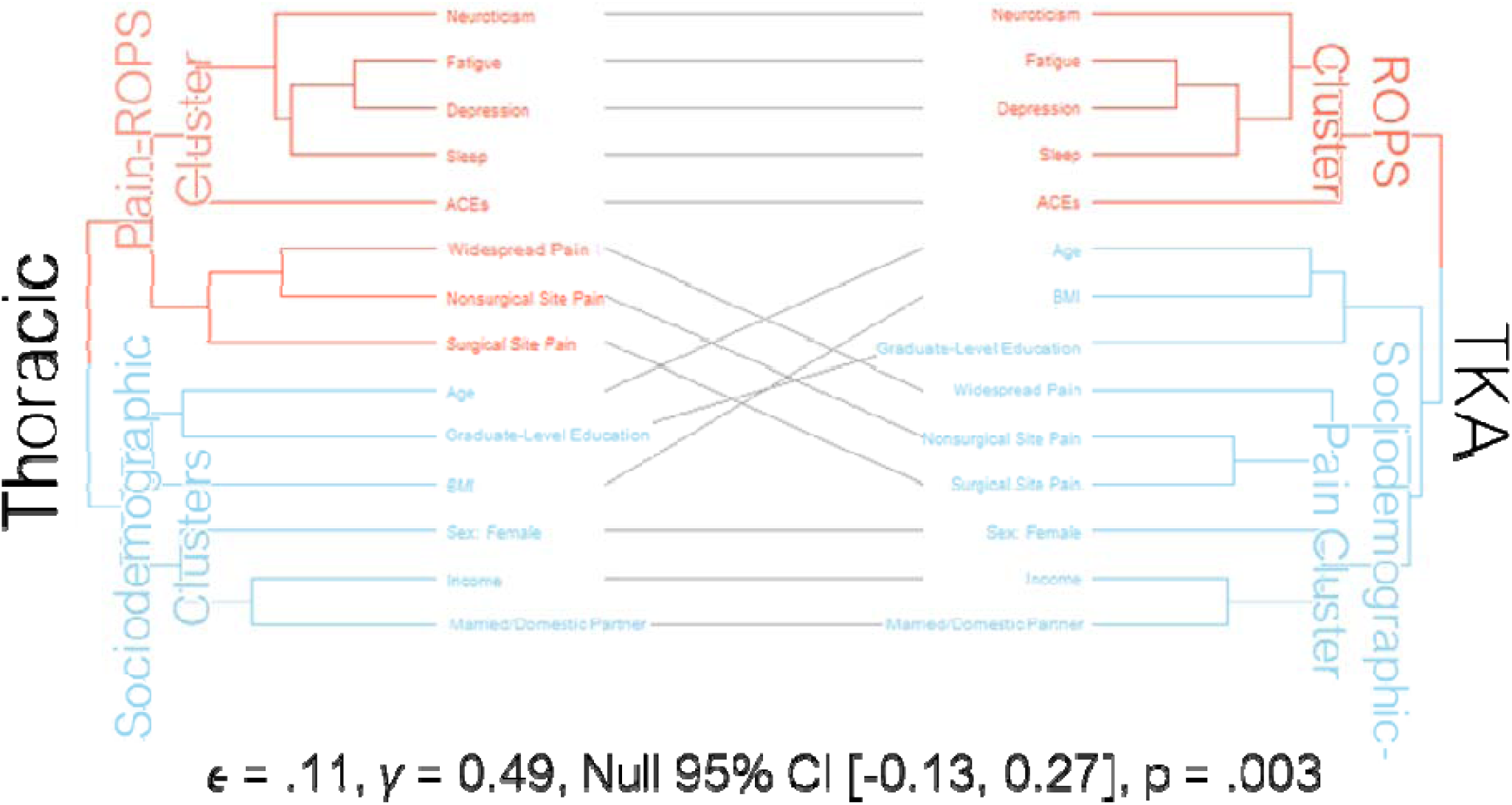
Tanglegram comparing correlational structure of pain and ROPS variables across cohorts. Comparison of cohort-specific clustering structure. A tanglegram contrasts the dendrograms derived separately for each cohort. Low entanglement (0.11) indicates high structural similarity between clustering solutions, and Baker’s gamma (0.49; p = 0.003) confirms that this similarity is unlikely to arise by chance. In the thoracic surgery cohort, pain outcomes clustered more closely with ROPS variables (excluding BMI), forming a unified cluster distinct from sociodemographic factors. In contrast, in the TKA cohort, ROPS variables (excluding BMI) clustered separately from pain outcomes.

### Single variable and multiple regression models

Having established that the latent ROPS framework generalized across cohorts, we examined which individual factors were independently associated with each pain outcome. In single-variable models, depression, fatigue, neuroticism, and sleep were each associated with all three pain outcomes in both cohorts, and ACEs with all three overall (though not with pain spread in the Thoracic cohort). BMI was the exception among the psychosocial factors: it predicted nonsurgical-site pain (+10%, p < .001) but not surgical-site pain or spread, and only within the TKA cohort (see Supplementary analysis). In multivariable models entering all factors simultaneously (Fig. 5), fatigue remained the only consistent independent correlate across outcomes (with household income the sole exception), identifying it as the dominant individual indicator and motivating the network and mediation analyses that follow.

**Figure 5.**
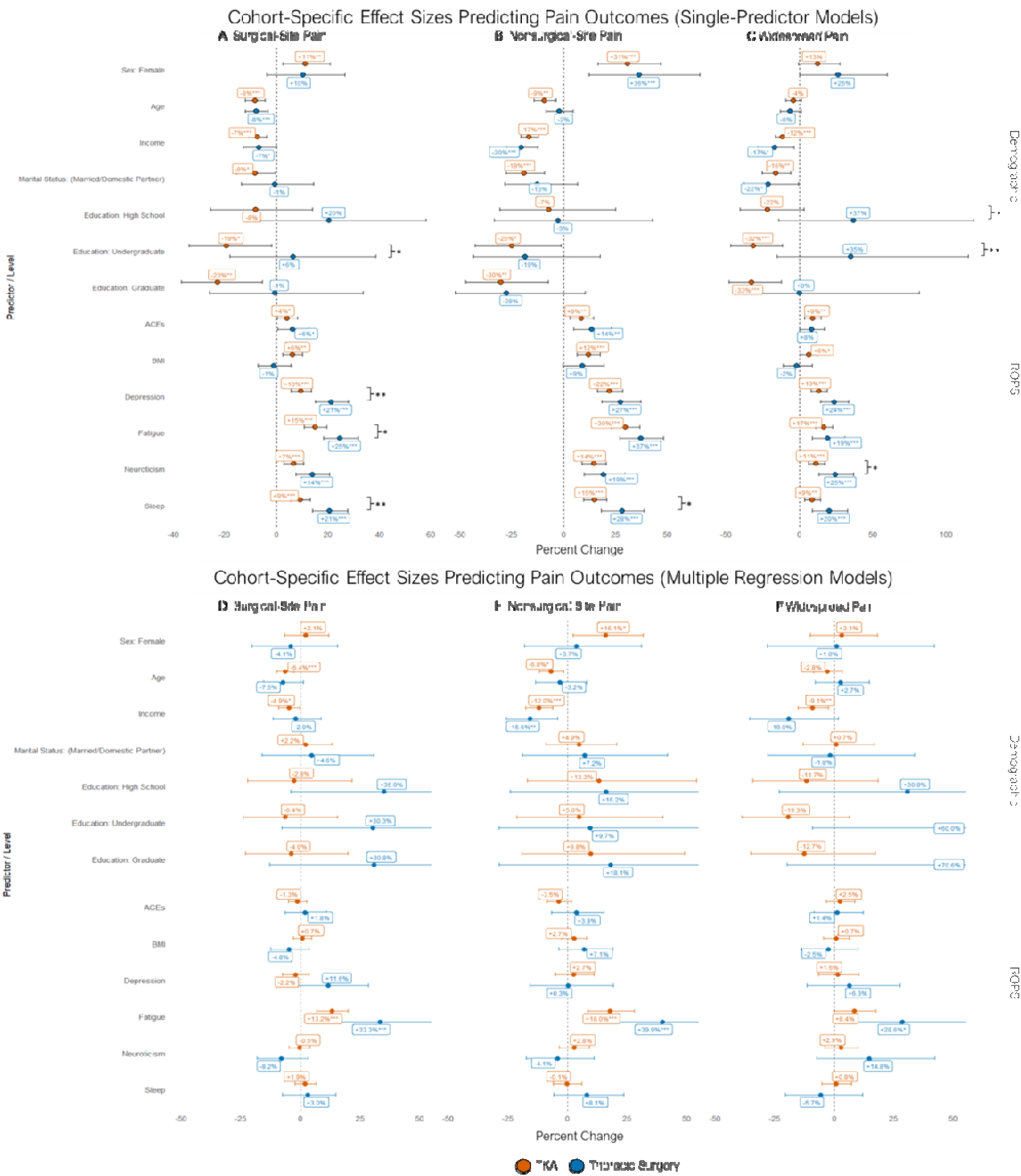
Univariate and multivariable predictors of presurgical pain outcomes by cohort. Comparison of single-variable and multivariable regression models predicting presurgical pain outcomes. Top panels show standardized effects from single-variable regression models estimated in the combined sample, with cohort-specific means overlaid Bottom panels show cohort-specific multivariable regression models including all predictors simultaneously. Models predict (A, D) surgical-site pain (SSP), (B, E) non-surgical-site pain (NSSP), and (C, F) widespread pain, estimated separately for the thoracic surgery and total knee arthroplasty (TKA) cohorts.

BMI was associated with pain only in the TKA cohort; however, the between-cohort difference was not statistically significant and the pooled estimate was null, so we do not interpret this as a cohort-specific effect. Female sex was associated with greater nonsurgical-site pain within each cohort separately and in the combined-cohort model adjusting for cohort and a cohort-by-sex interaction, indicating that this association is not attributable to the cohorts’ differing sex composition; it attenuated after adjustment for the other ROPS factors, remaining significant only for nonsurgical-site pain in the TKA cohort (Supplementary analyses). Full coefficients, 95% confidence intervals, and p-values for all models are provided in Table 2 (multivariable models) and Supplementary Table S2 (single-variable models).

**Table 2.** Multivariable associations of psychosocial and sociodemographic factors with presurgical pain outcomes, by cohort. Factors retained as independently associated in multivariable models entering all predictors simultaneously; %Δ = percent change in the (log-linked) outcome; VI = relative variable importance (0–100). Fatigue is the only factor independently associated with every outcome in both cohorts.

| Cohort | Outcome | Independently associated factors (%Δ [95% CI]; relative variable importance, VI) |
| --- | --- | --- |
| Thoracic | SSP | Fatigue +33.3 [14.5,55.1]*** (VI 100) |
| Thoracic | NSSP | Fatigue +39.9 [16.0,68.6]*** (VI 100); Household income -15.6 [-25.7,-4.1]* (VI 71) |
| Thoracic | Widespread | Fatigue +28.6 [4.5,58.4]* (VI 100) |
| TKA | SSP | Fatigue +13.2 [6.8,20.0]*** (VI 100); Age -6.4 [-10.0,-2.7]*** (VI 72); Household income -4.9 [-9.2,-0.3]* (VI 44) |
| TKA | NSSP | Fatigue +18.0 [8.7,28.0]*** (VI 100); Household income -12.0 [-17.5,-6.1]*** (VI 78); Female sex +16.1 [2.3,31.8]* (VI 50); Age -6.8 [-11.7,-1.7]* (VI 50); |
| TKA | Widespread | Household income -9.1 [-15.3,-2.4]** (VI 97); Fatigue +8.4 [-0.1,17.5]<sup>†</sup> (VI 84) |

<sup>†</sup> p < .10, \* p < 0.05, \*\* p < 0.01 and \*\*\* p < .001.

### Network analysis

We next asked how these variables were statistically organized by examining the conditional directional dependency structure underlying regression findings. To do this, we estimated Bayesian network (BN) structures separately for the Thoracic surgery and TKA cohorts as directed-acyclic graphs (DAGs). Across both cohorts, fatigue occupied the most central position in the Bayesian networks regardless of centrality metric (Fig. 7). Fatigue consistently linked upstream psychosocial and sociodemographic factors to downstream pain outcomes, whereas demographic variables occupied more peripheral positions.

In the Thoracic surgery DAG (Fig 6A), fatigue emerged as a central hub (Fig. 7A), receiving direct input from income, depression, and sleep quality, and in turn influencing SSP, NSSP, and spread. Fatigue and neuroticism were close to pain outcomes (∼0.18–0.55), while demographic predictors (sex, age, education, marital) were more distal. SSP and NSSP carried relatively high eigenvector centrality (structurally important sinks). Fatigue and Sleep Quality dominated authority scores. Neuroticism also served as a key connector. Sleep Quality mediated links from ACEs and Neuroticism to Depression and Fatigue, while Neuroticism mediated links from ACEs to BMI, Depression, Sleep Quality, and widespread pain. Pain outcomes clustered together, with directed connections from SSP, to NSSP, and lastly to spread.

**Figure 6.**
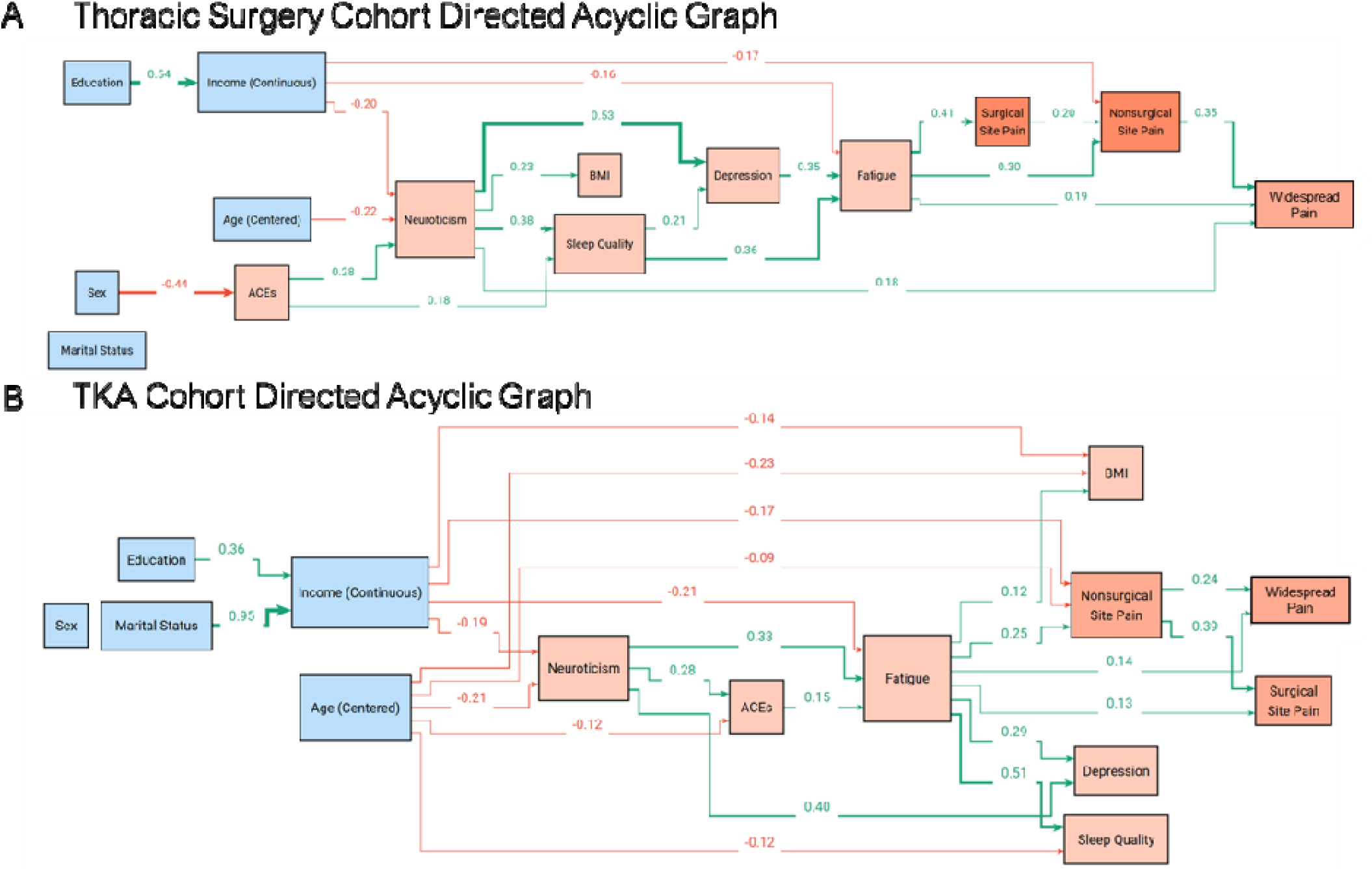
Fatigue-centered causal network structure of presurgical pain across cohorts. Directed acyclic graphs (DAGs) representing cohort-specific Bayesian network structures. Shown are DAGs estimated separately for the (A) thoracic surgery cohort and (B) total knee arthroplasty (TKA) cohort. Nodes represent sociodemographic (blue), Risk of Pain Spread (ROPS; light orange), and pain outcome (dark orange) variables; arrows indicate conditional dependencies after accounting for all other variables in the network. Across cohorts, fatigue emerged as a central hub linking upstream sociodemographic and psychosocial factors to pain outcomes. Pain outcomes—surgical-site pain (SSP), non-surgical-site pain (NSSP), and widespread pain—formed a tightly connected subnetwork, with cohort-specific differences in connectivity. The TKA network showed greater overall integration, including stronger involvement of BMI, neuroticism, and depression.

**Figure 7.**
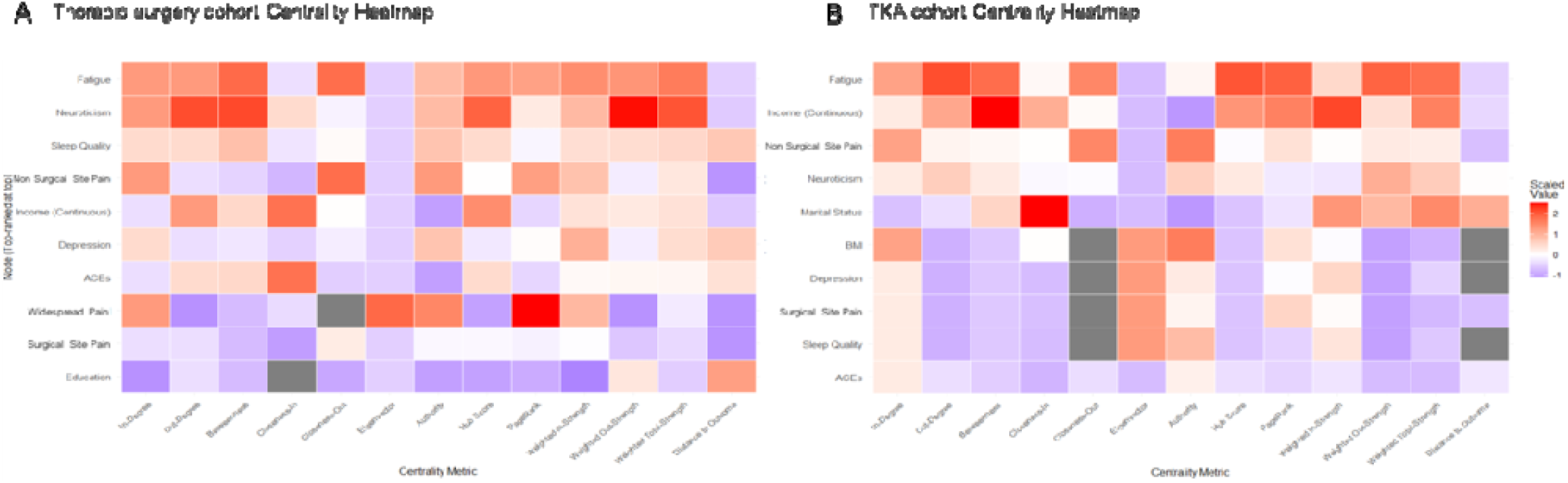
Fatigue is the most central node in cohort-specific pain networks. Centrality metrics for cohort-specific directed acyclic graphs. Heatmaps display the top 10 most central variables in the Bayesian network models for the (A) thoracic surgery cohort and (B) total knee arthroplasty (TKA) cohort. Centrality measures quantify each variable’s relative importance within the network structure. Across both cohorts, fatigue ranked as the most central indicating a prominent role in linking sociodemographic and psychosocial risk factors to pain outcomes.

In contrast, the TKA DAG (Fig. 6B) showed a more tightly integrated network. Fatigue again occupied a central position, dominating all metrics (pagerank, hub score, eigenvector; Fig. 7B) and directly predicting SSP and NSSP, and spread. Fatigue was predicted by ACEs, BMI, and neuroticism, and in turn predicted depression and sleep quality. NSSP was predicted by age and income as well, which in turn predicted SSP and spread. Income and marital status were surprisingly proximal as well (∼0.17–0.58). This suggests that socioeconomic variables are more entangled in the TKA rather than the Thoracic surgery cohort.

### Directionally-Constrained Model Comparison

We next compared competing hypotheses regarding statistical directionality by testing. w four directionally-constrained models that differed in how relations between fatigue and pain variables were specified. These models were designed to evaluate whether the observed covariation structure was more consistent with fatigue functioning as an upstream driver, a downstream consequence, or a mutually determined correlate of pain outcomes: (i) the original Fatigue→Pain [F2P] set, in which fatigue is treated as a proximal driver of pain, (ii) Pain→Fatigue [P2F] in which pain severity and pain widespread are assumed to influence fatigue and (iii) unconstrained [UNC] models, in which fatigue–pain relations are determined by structure learning without directional restrictions. We also tested (iv) a fatigue-as-outcome [FOUT] specification to evaluate whether the observed centrality of fatigue could be explained solely by its role as a downstream consequence rather than a driver within the network. Model comparison was done using the conditional Gaussian BIC (BIC-cg; higher values indicate better fit).

In the Thoracic surgery cohort, F2P and UNC were essentially tied (BIC-cg = −7065.445 for both; ΔBIC = 0), suggesting that the data do not meaningfully distinguish whether Fatigue→Pain constraints improve fit over the baseline constraints. P2F fit (ΔBIC = −0.040) and FOUT fit (ΔBIC = −1.70) was worse. In the TKA cohort, F2P also provided the best fit (BIC-cg = −22652.485), with UNC (ΔBIC = −0.61), P2F (ΔBIC = −5.24), and FOUT (ΔBIC = −20.80) being worse.

These findings indicate that models specifying fatigue as upstream of pain outcomes were more consistent with the observed covariance structure than competing alternatives. These findings indicate that modeling pain as a downstream outcome of fatigue—rather than modeling fatigue as downstream of pain or as the ultimate outcome—is more consistent with the observed covariation structure in both clinical cohorts.

Taken together, these networks highlight fatigue as a consistent central determinant across cohorts, bridging sociodemographic and psychosocial predictors with pain outcomes. Neuroticism and depression further reinforced these pathways, particularly in the TKA cohort where BMI also contributed. The cross-cohort comparison suggests that while fatigue universally links upstream vulnerability factors to pain, the embedding of BMI and mood symptoms into the pain network may be stronger in individuals undergoing TKA.

### Mediation Models

Finally, we tested whether complementary mediation analyses recapitulated the network structure. Mediation models informed by the F2P DAGS were tested to test indirect effects on pain outcomes, and the results are summarized in Table 3 Average Indirect Effects (AIE), Average Direct Effects (ADE), and Proportion Mediated (PM) were estimated using nonparametric bootstrapping. For interpretability, pathways were categorized as: full mediation (AIE significant; ADE not significant), partial (complementary) mediation (AIE and ADE significant and same sign), competitive (suppression) (AIE and ADE significant with opposite signs), direct-only (ADE significant; AIE not), total-only (TE significant; AIE and ADE not), or no effect (none significant). Because multiple, hypothesis-driven pathways were prespecified by DAGs, we emphasize effect sizes and interval estimates; no multiplicity correction was applied. Consistent with the Bayesian networks, fatigue emerged as the most frequent statistical mediator across both cohorts

**Table 3.**
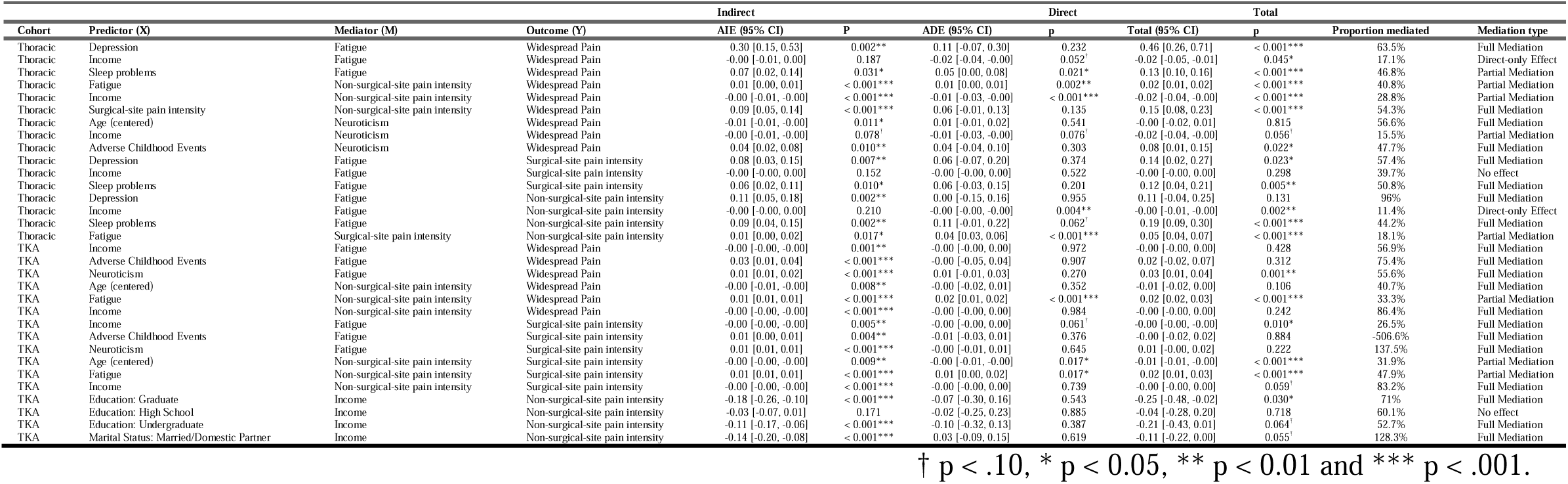
Directed acyclic graph–guided mediation analyses of presurgical pain outcomes. Shown are mediation models linking predictors (X) to pain outcomes (Y) through DAG-consistent mediators (M), estimated separately for thoracic surgery and total knee arthroplasty (TKA) cohorts. Indirect effects are reported as average indirect effects (AIE), direct effects as average direct effects (ADE), and total effects with 95% confidence intervals. Proportion mediated reflects the fraction of the total effect attributable to the indirect pathway. Mediation type indicates full, partial, or direct-only mediation based on the significance of indirect and direct paths. All models adjusted for cohort-specific covariate structure implied by the DAGs.

### Thoracic surgery cohort

Across pain outcomes for the thoracic surgery cohort, fatigue emerged as the most consistent upstream statistical mediator, particularly for depression and sleep problems. For SSP and NSSP, the associations of both depression and sleep problems with pain were largely or fully explained by indirect effects through fatigue, with negligible direct effects once fatigue was included (Table 2). Household income showed limited or inconsistent indirect effects through fatigue for SSP and NSSP, and in several cases operated primarily through direct or alternative pathways. For chronic widespread pain, both NSSP and fatigue served as key intermediaries. The effect of SSP on widespread pain was fully mediated by NSSP, whereas fatigue exerted both direct and indirect effects on spread through NSSP. Using fatigue as the mediator, depression showed a strong indirect association with widespread pain, whereas household income exhibited primarily direct effects. When neuroticism was modeled as the mediator, age and ACEs influenced widespread pain predominantly through neuroticism, while household income showed partial mediation. Taken together, the Thoracic cohort results indicate a multistep structure in which psychosocial vulnerability factors influence fatigue and internalizing ROPS traits, which in turn shape pain intensity and ultimately widespread pain.

### TKA cohort

In the TKA cohort, mediation patterns were more centralized around fatigue and NSSP. For SSP, the effects of age and fatigue were partially mediated by NSSP, whereas income effects were largely indirect. For both SSP and NSSP, income, neuroticism, and ACEs showed substantial indirect effects through fatigue, with weak or non-significant direct effects (Table 2). For NSSP, socioeconomic indicators—including marital status and educational attainment—were primarily associated with pain via income, which functioned as a key intermediary. For chronic widespread pain, NSSP fully mediated the effects of income and age, while fatigue showed both direct and indirect associations with spread. When fatigue was specified as the mediator, income, adverse childhood events, and neuroticism were predominantly linked to widespread pain through indirect pathways. Overall, the TKA cohort exhibited a more tightly integrated mediation structure, with fatigue and non-surgical pain acting as central conduits through which sociodemographic, and ROPS factors related to pain outcomes.

## Discussion

Across two large pre-surgical cohorts from the A2CPS Data Release 2.0.0, we externally validated the Risk of Pain Spread (ROPS) framework in clinically distinct surgical populations and identified fatigue as its most consistent and central correlate of pain burden. Despite substantial differences in age, sex, socioeconomic profile, pain burden, and surgical indication, the latent ROPS construct generalized across both cohorts and predicted surgical-site (SSP), nonsurgical-site (NSSP), and widespread pain. Across structural equation modeling, multivariable regression, Bayesian network, and mediation analyses, fatigue consistently emerged as the strongest independent correlate linking biopsychosocial vulnerability with pain intensity and spread, extending prior work implicating fatigue in centralized and widespread pain [5,6,16,30] and suggesting that it may be a proximal manifestation through which cumulative risk is expressed.

Although ROPS was derived to predict widespread pain, its factors were associated with all three pain outcomes in both cohorts. Because the framework was trained on widespread pain, it could in principle have related only to spread; instead it also tracked the intensity of localized pain, including surgical-site pain at a putatively nociceptive site (the osteoarthritic knee). This indicates that the vulnerability indexed by ROPS is relevant across pain types, not only widespread or centralized pain, and extends the framework beyond the community setting in which it was developed—UK Biobank lacked a validated, clinically defined nociceptive site of this kind. The latent construct was psychometrically stable across cohorts, but its associations with pain were consistently stronger in the Thoracic than the TKA cohort, particularly for widespread pain. One plausible explanation is that the substantial peripheral nociceptive drive of advanced knee osteoarthritis partially obscures the influence of underlying vulnerability, whereas pain in Thoracic participants may more directly reflect centrally mediated processes—consistent with evidence that the relative contributions of nociceptive and centralized mechanisms vary across conditions [2,8,18]. Regardless, fatigue remained the most consistent correlate in both cohorts.

Nociplastic contributions are plausible across all three outcomes. NSSP and widespread pain are conceptually closer to centralized/nociplastic pain, whereas surgical-site pain is often assumed to be primarily nociceptive; yet ROPS predicted all three, and the outcomes were themselves positively correlated. Associations were numerically somewhat larger for NSSP and widespread pain than for SSP, but the differences were small, and we do not interpret them as evidence that ROPS preferentially predicts nociplastic outcomes—particularly because widespread pain is a count without magnitude information and is not directly comparable to the intensity outcomes, and we did not formally test the difference in effect sizes.

Across regression, network, and mediation analyses, the statistical organization was most consistent with a two-stage pathway: upstream psychosocial load (socioeconomic disadvantage, adverse childhood experiences, neuroticism, depression, sleep disturbance) converged on fatigue; fatigue was closely associated with both SSP and NSSP; and NSSP served as a proximal correlate of widespread pain in both cohorts [2,8,15,19]. Structural equation modeling showed the latent ROPS construct predicted all three outcomes; multivariable regression identified fatigue as the most consistent independent correlate after adjustment; Bayesian networks positioned fatigue as a central hub with demographic factors more peripheral; and mediation analyses recapitulated the network-implied paths. Because the analyses are cross-sectional, these pathways reflect statistical organization rather than causal direction; forthcoming postoperative A2CPS data will allow prospective evaluation.

Network topology nonetheless differed across populations. The Thoracic cohort showed a relatively distributed structure, with fatigue, sleep disturbance, neuroticism, and depression all moderately central, whereas the TKA cohort was more centralized around fatigue, with household income in a more proximal position. Thus, while fatigue consistently linked upstream vulnerability to pain across contexts, the surrounding psychosocial architecture varied with pain etiology and clinical population.

Socioeconomic context was also important: lower household income was consistently associated with greater nonsurgical and widespread pain, directly and through fatigue and NSSP; younger age predicted greater pain in both cohorts; and female sex was associated primarily with greater nonsurgical pain, consistent with broader evidence that women more frequently report widespread pain and can face distinct challenges in having their pain clinically recognized [32]. These patterns reinforce that vulnerability to widespread pain reflects interacting biological, psychological, and socioeconomic factors rather than any single mechanism [20,30].

Several clinical implications follow. Fatigue should be routinely assessed as a potentially modifiable feature of chronic pain; given its centrality, interventions targeting fatigue—treating sleep disorders, optimizing mood, graded activity, and addressing metabolic contributors—may yield broad improvements [5–7,13]. Nonsurgical-site pain should be monitored across perioperative recovery, as it consistently occupied the position immediately preceding widespread pain and may flag patients at elevated risk [2,31]. Socioeconomic factors warrant consideration alongside biomedical risk, integrating social-risk screening and support into perioperative care [11,16].

Our findings extend and refine the original ROPS framework [30]. Whereas Tanguay-Sabourin and colleagues used dichotomized factors in a community cohort, we used continuous, validated measures in two clinically distinct surgical populations, allowing us to characterize not only whether the factors generalized but how they were organized: rather than contributing independently, they formed an interconnected network centered on fatigue. We also operationalized developmental adversity through adverse childhood experiences rather than contemporaneous stress; incorporating current life stress will further clarify how proximal and distal stressors contribute to spread.

Strengths of this work include two large, independently recruited surgical cohorts with harmonized phenotyping (BPI-SF, Michigan Body Map), theory-guided directed acyclic graphs constraining the network and mediation models, and convergent evidence across complementary analyses. Several limitations should be acknowledged. The design is observational and cross-sectional and cannot establish causation; causal interpretation of the mediation models rests on assumptions that cannot be verified with cross-sectional data. Because widespread pain is defined by pain at multiple sites, some association with NSSP is expected, though NSSP’s consistent emergence across methods suggests it reflects more than definitional overlap. Several constructs (fatigue, sleep, mood) were self-reported and subject to measurement error, missing data were handled by listwise deletion, and proportion-mediated estimates are scale-dependent under log links. A causal test would require interventions that reduce fatigue through non-nociceptive means; future work should extend ROPS to other clinical pain conditions and incorporate postoperative follow-up, objective sleep and activity monitoring, inflammatory and metabolic biomarkers, and fatigue-targeted trials [8,33].

In conclusion, we externally validated the ROPS framework in two clinically distinct pre-surgical cohorts and found fatigue to be the central feature linking biopsychosocial vulnerability with pain intensity and widespread pain. Although network structure differed across populations, the pattern was consistent: fatigue occupied a central position connecting upstream vulnerability with pain outcomes, while increasing nonsurgical-site pain closely accompanied widespread pain. These results clarify how multiple risk factors integrate into clinical pain burden and identify fatigue as a promising modifiable target for future mechanistic and interventional work aimed at preventing the transition from localized to widespread pain.

## Supporting information

Supplementary Analyses

## Data Availability

All data provided from the A2CPS consortium are available to other investigators online upon permission granted by https://a2cps.org/researchers/accessing-our-data/. Detailed code and annotation will be made publicly available at GitHub (https://github.com/canlab and https://github.com/a2cps).

## Acknowledgements

The authors acknowledge the contributions of members of the Acute to Chronic Pain Signatures (A2CPS) Consortium, including: Kumari Adams, Ambar Akhlas, Oluwaseyi “Seyi” Akintoroye, Raed Alnajjar, Miguel Alvelo-Rivera, Miracle Anderson, Aravind Athivirahham, Aishat Bakare, Madhumitha Balaji, Tessa Balach, Kortney Barrett, Emine Bayman, Rachel Bergmans, Seth Berke, Emby Black, Angela Broski (Polanco), Derrick Brown, Darren Bryan, John Burns, Asokumar Buvanendran, Brian Caffo, Vince Calhoun, James Carson, Anelizze Castro-Martinez, Michael Charters, David Chesla, Joe Chue, Laura Cin, Chris Coffey, Courtney Cole, Douglas Colquhoun, Danneka Cooper, Laura Cox, Michele Costigan, Ciprian Crainiceanu, Dana L. Dailey, Elizabeth Dailey, Mark Delano, Olivia Dionisio, Luda Diatchenko, Kendall Dubois, Nina Duong, Amy Durkin, Dixie J. Ecklund, Maximillian Egan, Farzad Farahani, Sarah Fazal, Oliver Fiehn, Katie Fisch, Michael Flannery, James Ford, Stephan Frangakis, Lars Fritsch, Laura Frey Law, Kara Friedman (Schoenberg), Ramtilak Gattu, Vennela Gajjala, Hagai Ganin, Karla Gendler, Sophie George, Adi Gherman, Elizabeth Goetz, Rachel Gorre, Emma Griebenow, Xiaodong Guo, Andre Hackman, Abbas Hakim, DeAnna Hanewald, Steven Harte, Kasper Hansen, Khaled Hasan, Emre Berk Hayir, Esmeralda Hidalgo-Lopez, Candy Hodges, Timothy Howard, Trevis Huff, Eric Ichesco, Jimmy Jagan, Adam Janowski, Kristen Jepsen, Hongkai Ji, Micah Johnson, Tanja Jovanovic, Ari B. Kahn, Muge Karaman, Olivia Keaveny, Sachin Kheterpal, Tobias Kind, Donald Koehler, Nivya Kolli, Mohan Kulkarni, Tony Larkin, Carl Langefeld, Angel Ledesma, Zackary Lemka, Nondas Leloudas, Remy Lobo, Anna Lokshin, Hue Luu, Qingfei Luo, Michael Liptay, Vincent A. Magnotta, John Mahajan, Maria Lucia Madariaga, Maggie Makar, Silvia Marroquin, Cayla Mason, Robert McCarthy, Sarah Meehan, Zaki Mehkri, Nathaniel Mendoza, Todd Mulderink, Leigh Nadel, Lillian (Anna) Nefcy, Tina Neill-Hudson, Kasia Nowak, Ikenna Okereke, Michael Olivier, Tyler Ostrander, Marc Parisien, Allison Pekar, Scott Peltier, Lissa Pearson, Jennifer Pierce, Hristo Piponov, Samantha Pianga (Law), Andrew Popoff, Thomas Prince, Pottumarthi Prasad, Hedda Prochaska, Sobha Puppala, Wei-Jun Qian, Laura Quigley, Ellen Quillen, Lynn A. Rasmussen, Chloe Reeves, Arisbeth Reyes, Ingo Ruczinski, Patrick Sadil, Pat Scherer, Andrew Schrepf, Jacey Schickel, Kathy Scott, Sydney Shohan, Christopher Sica, Michael Sipe, Anik Sinha, Maggie Spencer, Alexis Stanczuk, Stephani Sutherland, Alicia Suydam, Margaret Taub, Opal Tafe, Carla Tisdale, Joshua Urrutia, Andrew Urquhart, Shaurel Valbrun, Carol G. T. Vance, Matthew Vaughn, Pablo Vicente, Sara Wallace, Noah Waller, Benlian Wang, Jennifer Waljee, Sydney Whack, Amber White (DeGraaf), David Williams, Brian Winner, Sterling Winters, Denise Wittenbach, Melanie Wong, Yang Xuan, Ezgi Yarasir, Scott Zeger, Lara Zador, Yong Zhou, Joe Zhou, Ping-Shou Zhong, Guohao (Arthur) Zhu, and Katerina Zorina-Lichtenwalter.

