## Supplementary Analyses for "Fatigue Links Sociodemographic Risk to Pain Intensity and Spread in Two Surgical Cohorts"

#### *Correlogram*

Pairwise correlations among predictors and outcomes were computed using Spearman’s coefficients and visualized with correlograms. Hierarchical clustering was applied to correlation matrices, and dendrograms were compared across methods using tanglegrams to evaluate cluster stability. Correlograms were generated separately for each cohort and variables were clustered using Ward’s minimum variance method (Ward.D2). To compare the resulting dendrograms across cohorts, we used tanglegrams, where colors indicated cluster membership. We computed two complementary metrics: entanglement, which quantifies how well two trees can be aligned without crossing edges (0 = identical, 1 = maximally tangled), and Baker’s γ correlation, which assesses concordance of ultrametric distances between the trees (−1 to 1). Significance of Baker’s γ was evaluated using 5,000 permutations of the dendrogram labels to generate a null distribution.

A bivariate Spearman's correlogram (Supplementary Fig. S1) was computed separately for each cohort. Across both cohorts, female sex was positively correlated with NSSP, whereas age was negatively correlated with SSP in both cohorts and with NSSP in the TKA cohort. Lower household income was inversely correlated with multiple pain outcomes in the TKA cohort, and unmarried or unpartnered status was positively correlated with NSSP and widespread pain in the TKA cohort. Depression, neuroticism, sleep disturbance, and fatigue formed a robust cluster of positively correlated psychosocial factors that was broadly associated with pain outcomes, particularly in the TKA cohort. Fatigue exhibited the most consistent positive correlations with SSP, NSSP, and widespread pain across both cohorts. ACEs and BMI were positively correlated with NSSP and widespread pain in the TKA cohort but not in the Thoracic surgery cohort. Finally, SSP and NSSP were positively correlated in both cohorts, as were NSSP and widespread pain, whereas SSP was significantly correlated with widespread pain only in the Thoracic surgery cohort.

### *Single variable regression model main effects*

In cohort-specific single-variable regressions, female sex was associated with greater nonsurgical-site pain in both cohorts (Thoracic +36% [+12, +66], p < .001, R² = 0.034; TKA +31% [+17, +47], p < .001, R² = 0.029) and with greater surgical-site pain in the TKA cohort (+11% [+2, +21], p = .009, R² = 0.011), with weaker, marginal associations with widespread pain; it was not associated with surgical-site pain in the Thoracic cohort. Because these associations hold within each cohort analyzed separately, they cannot be attributed to the cohorts' differing sex composition. Consistent with this, in the combined-cohort model—which adds cohort and a cohort-by-sex interaction, so the sex main effect is averaged across cohorts—female sex was associated with greater nonsurgical-site pain (+33% [+19%, +49%], p < .001, partial R² = 0.030) and widespread pain (+19% [+5%, +36%], p = .008, partial R² = 0.003). These sex associations attenuated when the other ROPS factors were added, remaining significant only for nonsurgical-site pain in the TKA cohort, consistent with shared variance among the psychosocial factors.

Averaged across cohorts, greater surgical-site pain (Fig. 5A) was associated with younger age (−8% [−11%, −5%], p < .001, partial R² = 0.022), lower income (−7% [−11%, −3%], p < .001, partial R² = 0.017), and higher levels of each ROPS factor—ACEs (+5% [+2%, +9%], p = .004, partial R² = 0.007), depression (+15% [+12%, +19%], partial R² = 0.048), fatigue (+20% [+16%, +24%], partial R² = 0.076), neuroticism (+10% [+6%, +14%], partial R² = 0.021), and sleep problems (+15% [+11%, +19%], partial R² = 0.042), all p < .001. Greater nonsurgical-site pain (Fig. 5B) and greater pain spread (Fig. 5C) were additionally associated with female sex (nonsurgical-site +33% [+19%, +49%], partial R² = 0.030; spread +19% [+5%, +36%], p = .008, partial R² = 0.003). Higher education was associated with less nonsurgical-site pain (partial R² = 0.023): relative to the lowest-education group, undergraduate −22% [−38%, −2%], p = .030, and graduate −29% [−45%, −8%], p = .004. Being married or domestically partnered was associated with less widespread pain than those who were not (−19% [−29%, −8%], p < .001, partial R² = 0.006).

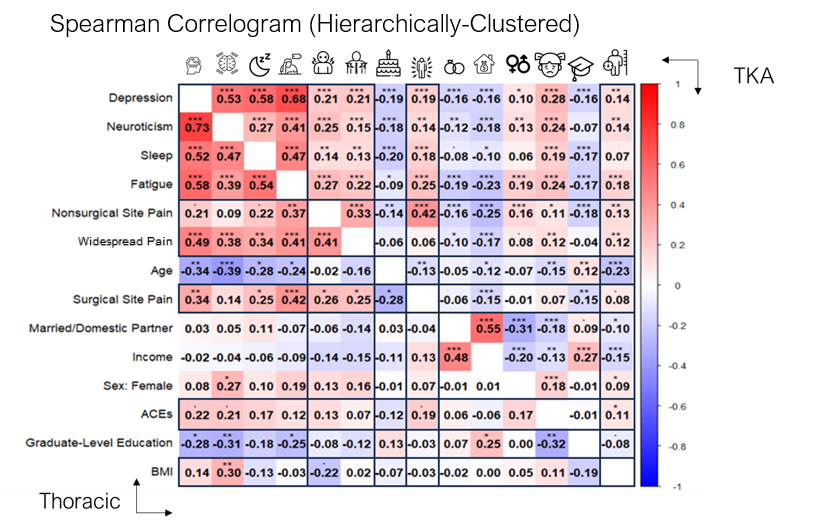
**Supplementary Figure S1. Spearman correlogram of pain and ROPS variables across cohorts.** Spearman correlation structure by cohort. Spearman correlogram showing the thoracic surgery cohort (lower-left triangle) and the total knee arthroplasty (TKA) cohort (upper-right triangle). Variables were hierarchically clustered based on the average correlation structure across cohorts, revealing close overall proximity between Risk of Pain Spread (ROPS) variables and pain outcomes.

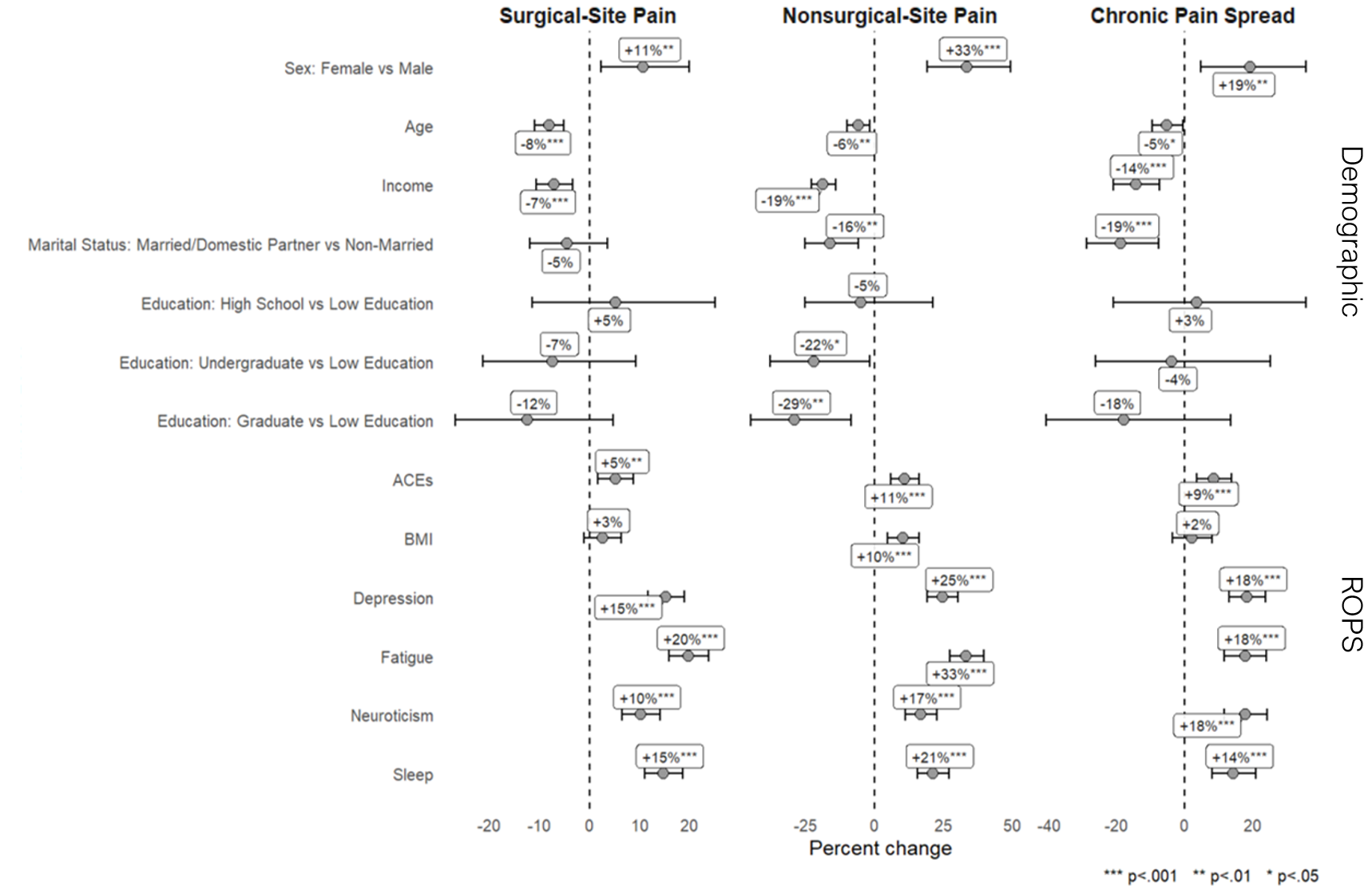

**Supplementary Figure S2.** Single-variable associations of ROPS predictors with presurgical pain outcomes. Standardized effects from single-variable regression models predicting presurgical pain outcomes. Shown are standardized regression coefficients for Risk of Pain Spread (ROPS) predictors and sociodemographic variables predicting (A) surgical-site pain (SSP), (B) non-surgical-site pain (NSSP), and (C) widespread pain. Across outcomes, greater pain severity was associated with higher fatigue, depressive symptoms, adverse childhood experiences (ACEs), neuroticism, and sleep disturbance, as well as lower household income and younger age. Female sex showed stronger associations with NSSP and widespread pain, and lower educational attainment was selectively associated with higher NSSP.
